# Data sharing in cancer research: A qualitative study exploring community members’ preferences

**DOI:** 10.1101/2024.07.21.24310665

**Authors:** Elizabeth A. Johnston, Xanthia E. Bourdaniotis, Susannah K. Ayre, Leah Zajdlewicz, Vanessa L. Beesley, Belinda C. Goodwin

## Abstract

Advancements in cancer treatment and survivorship rely on participation in research and access to health records. This study explored preferences for data access and sharing in 14 workshops with 42 community members, most of whom were a cancer survivor or carer. Various scenarios for data access and sharing were presented and discussed, with participants’ preferences summarized using descriptive statistics. Reasons underlying these preferences were identified through a thematic analysis of workshop transcripts. Most participants indicated a willingness for researchers to use their self-reported data and current health records for a specific research project (86%). Many were also willing for their self-reported data and current (62%) or all future (44%) heath records to be shared with other researchers for use in other studies if made aware of this. Willingness to consent to data access and sharing data in cancer research was influenced by: (i) the potential for data sharing to advance medical discoveries and benefit people impacted by cancer in the future, (ii) transparency around researchers’ credibility and their intentions for data sharing, (iii) level of ownership and control over data sharing, and (iv) protocols for privacy and confidentiality in data sharing. Based on these themes, we present practical strategies for optimizing data access and sharing in cancer research.

## Introduction

With cancer a leading cause of mortality worldwide (1), there is an ongoing need for research-driven innovations in cancer treatment and survivorship (2). Achieving this requires researchers to collect, store, and analyze a wide range of data, including demographic, health, psychosocial, behavioral, and genetic information (3). Collecting these data from a large, representative samples in target populations is important for generating valid research outputs that can inform future interventions (4). Further, using these data in other research projects or sharing with other researchers reduces participant burden, prevents duplication of work, and supports mutual resource use (5). Thus, there is a need to not only optimize community participation in cancer research, but also facilitate efficient data collection and sharing.

The collection, use, storage, and sharing of human research data is tightly regulated through legislative and institutional policies and guidelines to protect the privacy and well-being of research participants. For example, in Australia, researchers are required to adhere to the principles and guidelines outlined in the National Statement on Ethical Conduct in Human Research in order to receive ethical approval and funding from research institutions and funding bodies (6). In most instances, these guidelines require researchers to disclose information about the purpose of the research to prospective participants, including what their participation will involve, and obtain their voluntary consent to participate, which if feasible, may be withdrawn at any stage (6). Ethical approval also requires that personal information is stored securely and kept confidential (6). Such regulation serves to protect individuals with respect to the use of their data but can be perceived as a barrier to processes such as data sharing and linkage (7).

When consumers are considering taking part in medical research, review evidence indicates that a key determinant of research participation is how their data will be collected, stored, and accessed (8). Some studies have investigated consumers’ preferences for data sharing within the oncology setting, particularly in the United States, identifying a willingness for data sharing beyond the immediate research project but a need for further disclosure around sharing practices (9–11). However, little is known about the principles that underly consumers’ preferences for how their data are used in cancer research and how studies could be designed to optimize data collection and sharing. This paper explores community members’ preferences for use of their self-report data and health records in cancer research. It also examines the reasons underlying consumers’ preferences for data access and sharing in cancer research and provides practical recommendations for designing and conducting cancer research studies. Understanding and applying consumers’ preferences for data access and sharing will support research integrity and may optimise community participation in cancer research.

## Methods

### Context and setting

Data were collected for this study during workshops held to co-design a population-based cancer survivorship study in Queensland, Australia, with methods described elsewhere (12). Briefly, co-design research actively involves consumers in developing, designing, implementing, and evaluating new products or services (13). Consumer involvement in research can contribute to better study outcomes, including higher enrolment and retention rates (14). The current study adopted a pragmatic approach to co-design with community members participating in workshops to design study invitation materials and a qualitative survey tool for collecting detailed information on the supportive care needs and experiences of people affected by cancer (12). The session activities and visual materials used in the workshops were developed by the broader study team, including cancer survivorship researchers, medical oncologists, and executive and senior leaders in a cancer support organisation and a population-based registry. This paper reports on findings from the final activity in the workshop that was completed in 14 of the 15 workshops (*n*=42 community members from a total sample of 44) (see **Figure 1**). Ethical approval for the study was obtained from the University of Southern Queensland Human Research Ethics Committee (ref: ETH2023-0140).

**Figure 1.**
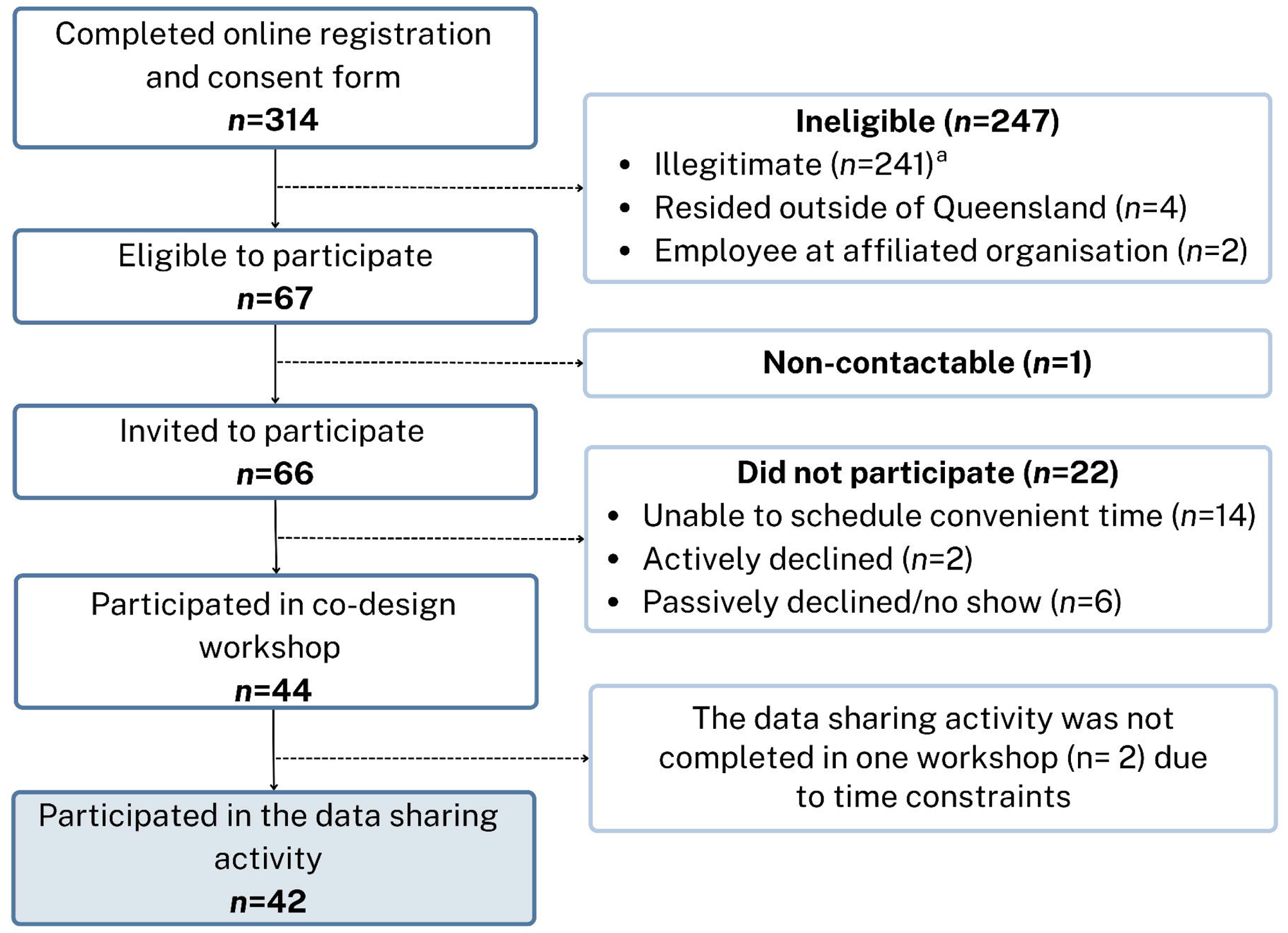
Flowchart of participant recruitment and selection for the co-design workshops. ^a^ Invalid/ineligible responses to the workshop invitation were identified based on a combination of factors (e.g., in the past duplicated IP addresses with different names, invalid postcodes or phone numbers, replicated responses in a short period of time, unusual completion times have been indicative of phishing attempts). Where validity could not be determined from the response, it was investigated further through phone and/or email contact with the respondent.

### Participants and recruitment

Participant recruitment was undertaken from October to December 2023 using digital and printed flyers distributed via networks associated with Cancer Council Queensland or the broader research team. To support the recruitment of priority populations, such as culturally and/or linguistically diverse (CALD) groups, the research team submitted study information to a health consumers network for inclusion in their e-newsletter (15). As data collection advanced, recruitment was supplemented through snowball sampling, with workshop participants invited to share recruitment flyers with friends, family members, and colleagues. Community members were eligible to participate in the co-design workshops if they were aged 18 years or older, English-speaking, and residing in Queensland, Australia. Participants included people with a personal experience of cancer (either as someone diagnosed with cancer or a carer for someone with cancer), as well as community members who did not have a personal experience of cancer. The latter group was included as current cancer incidence rates means it is likely that everyone will be impacted by cancer at some stage in their life, either as a patient or carer (16).

Participants were allocated to a workshop based on their nominated availability, with no more than five registered participants per workshop. Recruitment continued until a diverse sample had been achieved and the research questions had been adequately explored, determined by the authors through concurrent data collection and analysis. Due to the large number of online registrations for the interviews, participants were purposively sampled based on their demographic characteristics, including gender, ethnicity, Indigenous status, and geographical location, as well as their experience with cancer (i.e., survivor or carer) to ensure that diverse perspectives were represented.

### Data collection

Workshops were facilitated by two female researchers with undergraduate or postgraduate degrees in health science fields and training in qualitative data collection. The facilitators had no prior relationship with workshop participants. At the start of each workshop, the facilitators introduced themselves, including their role in the research team and their academic background. Workshops were conducted as either online (n=9), in-person (n=1), or hybrid (i.e., online and in-person) (n=4) sessions using Microsoft Teams. In-person participants attended the session at one of two not-for-profit organisations, where participants were provided with the relevant materials (e.g., pen, paper). Participants attending online were asked to source these materials themselves Workshops were audio-recorded and transcribed using Microsoft Teams. After completing the workshop, participants received a voucher valued at AU$100.00 for their time (approx. 120 minutes). Data for the current study were drawn from the final activity of the workshops, when participants were given a series of hypothetical scenarios developed by the research team to prompt discussion about preferences for data collection and sharing in cancer research (**Table 1**). Participants were advised that in the scenarios, ‘self-report data’ refers to information about themselves that they provide to researchers (e.g., needs and experiences), while ‘health records’ refers to information about themselves that researchers gather from registries or medical documents (e.g., cancer type and stage). The facilitator then provided a short explanation for each scenario (see italics in **Table 1**). For scenarios that involved data sharing, participants were informed that their information would be kept confidential. The lead facilitator asked each participant to verbally indicate which scenario (i.e., level of data sharing) they felt comfortable with and discuss the reasoning behind their choice. This often resulted in further discussion among group members around data sharing in cancer research.

**Table 1.** Preferences for data access and sharing in cancer research among community members participating in workshops to co-design a population-based cancer research study.

| Scenarios used to prompt discussion about use of data in cancer research<sup>†</sup> | Total<br>N=42 | Personal experience of cancer |  | No personal<br>experience of cancer<br>N=8 |
| --- | --- | --- | --- | --- |
|  |  | Diagnosis<br>N=18 | Carer<br>N=16 |  |
| Where do you sit on the scale? | N (%) | N (%) | N (%) | N (%) |
| 1. I am <u>okay</u> with self-report data that I provide directly to researchers being used for a specific research project. *This is the same as what you consented to for this workshop.* | **42 (100%)** | 18 (100%) | 16 (100%) | 8 (100%) |
| 2. I am <u>okay</u> with #1 + my current health records being used for a specific research project. *This means you would not have to self-report information that can be accessed directly from your health record, such as cancer type and stage.* | **36 (86%)** | 14 (78%) | 15 (94%) | 7 (88%) |
| 3. I am <u>okay</u> with #2 + my self-report data and my current health records being shared with other researchers for other projects if I am made aware of them. *Your information would be kept confidential, but your information could be used to provide data for other projects that you are made aware of.* | **26 (62%)** | 10 (56%) | 11 (69%) | 5 (63%) |
| 4. I am <u>okay</u> with #3 + future health records being shared with other researchers for other projects if I am made aware of them. *Similar to the previous option, but you are providing consent for ongoing collection of information about you from your health record that you will be made aware of.* | **18 (43%)** | 6 (33%) | 7 (44%) | 5 (63%) |

<sup>†</sup> Scenarios are not mutually exclusive and build on each other (i.e., participants who selected Scenario 3 were also comfortable with Scenarios 1 and 2).

### Data analysis

Descriptive statistics were used to summarize participant characteristics and preferences for data access and sharing based on the scenarios presented. Workshop transcripts were analyzed using codebook thematic analysis to identify recurring patterns in the data. As described by Braun and Clarke, codebook thematic analysis is a structured approach to coding that conceptualises themes as topic summaries of a central concept and is distinct from their reflexive approach to thematic analysis (17). First, two members of the research team (XB, EJ) reviewed workshop transcripts to familiarise themselves with the data. Second, transcripts were coded inductively by one researcher (XB) based on the words used by participants to describe their data sharing preferences and reasons underlying their preferences. These codes were documented in a coding framework alongside representative participant quotes. Third, initial themes were generated by grouping codes together that addressed the same concept (e.g., the need for privacy and confidentiality). Within each theme, initial sub-themes were developed to capture the various perspectives and nuances expressed by participants for each concept. Fourth, themes and sub-themes were further developed through discussion with a second researcher (EJ) and documented using an audit trail. Finally, each theme was assigned a title to summarize the key concept that it represented.

## Results

The characteristics of the 42 workshop participants are shown in **Table 2**. Participants had a median age of 43 years (range 23-79 years) and 31 (74%) were female. Thirty-four (81%) people had a personal experience of cancer, either as someone diagnosed with cancer (*n*=18; 43%) or as a carer of someone diagnosed with cancer (*n*=16; 38%). Eleven (26%) people were living in a regional or remote area and 19 (45%) lived in an area of low to medium socioeconomic status. Nine (21%) people were born overseas, 4 (10%) spoke a language other than English at home, and 3 (7%) identified as Aboriginal and/or Torres Strait Islander.

**Table 2.** Characteristics of participants in the 14 co-design workshops (n=42)

| Characteristic | Co-design workshops, N (%)<sup>a</sup> |
| --- | --- |
| **Age (years)** |  |
| Median (range) | 43 (23-79) |
| **Gender** |  |
| Female | 31 (74%) |
| Male | 11 (26%) |
| **Born overseas** |  |
| No | 31 (74%) |
| Yes | 9 (21%) |
| Not reported | 2 (5%) |
| **Ethnicity** |  |
| Caucasian | 34 (81%) |
| Asian | 5 (12%) |
| Aboriginal and/or Torres Strait Islander | 3 (7%) |
| **Language spoken at home** |  |
| English only | 36 (86%) |
| Other | 4 (9%) |
| Unknown | 2 (5%) |
| **Geographic remoteness (ARIA)** |  |
| Major city | 31 (74%) |
| Regional or remote | 11 (26%) |
| **Area-level socioeconomic status (SEIFA)<sup>b</sup>** |  |
| High | 23 (55%) |
| Medium | 13 (31%) |
| Low | 6 (14%) |
| **Personal experience with cancer** |  |
| Cancer survivor | 16 (38%) |
| Cancer caregiver | 16 (38%) |
| Both | 2 (5%) |
| Neither | 8 (19%) |
| **Time since diagnosis** |  |
| <1 year | 4 (10%) |
| 1-5 years | 6 (14%) |
| >5 years | 7 (17%) |
| Unknown | 1 (2%) |
| Not applicable | 24 (57%) |
| **Duration of caregiving** |  |
| <1 year | 7 (17%) |
| 1-5 years | 5 (12%) |
| >5 years | 5 (12%) |
| Unknown | 1 (2%) |
| Not applicable | 24 (57%) |

**Abbreviations:** ARIA, Accessibility/Remoteness Index of Australia; SEIFA, Socio-Economic Indexes for Areas
<sup>a</sup> Number of participants and percentage of sample unless otherwise stated.
<sup>b</sup> Higher scores indicate higher relative socioeconomic advantage and lower relative socioeconomic disadvantage in general. High = deciles 7-10, medium = deciles 4-6, low = deciles 1-3.

### Preferences for data access and sharing in cancer research

Preferences for data access and sharing in cancer research are shown in **Table 1**. All participants agreed with the baseline scenario of providing self-report data directly to researchers for a specific research project, as they had done for the workshops reported in this paper. In addition to providing self-report data, thirty-six (86%) would be willing to grant researchers access to their *current* health records for a specific research project. Twenty-six (62%) would be willing for their de-identified data and *current* health records from the original research project to be shared with other researchers for other projects if they were made aware of them. Less than half (n=18; 44%) would be willing for their de-identified *future* health records to be shared with other researchers for other projects if they were made aware of them. Of the 26 people who would be willing for their self-report data to be shared with other researchers for other projects, 5 participants (19%) felt they would not need to be made aware of how their information was being used (i.e., receive information about the other research projects and other researchers). These participants included 3 cancer survivors, 1 cancer carer, and 1 person with no personal history of cancer.

### Reasons underlying preferences for data access and sharing in cancer research

Four themes were identified, representing key principles that underpinned participants’ willingness to share their self-report data and health records in cancer research. These themes were: (i) the potential for data sharing to advance cancer research and benefit people impacted by cancer in the future, (ii) transparency around researchers’ credibility and their intentions for data sharing, (iii) level of ownership and control over data sharing, and (iv) protocols for privacy and confidentiality **(Table 3)**.

**Table 3.**
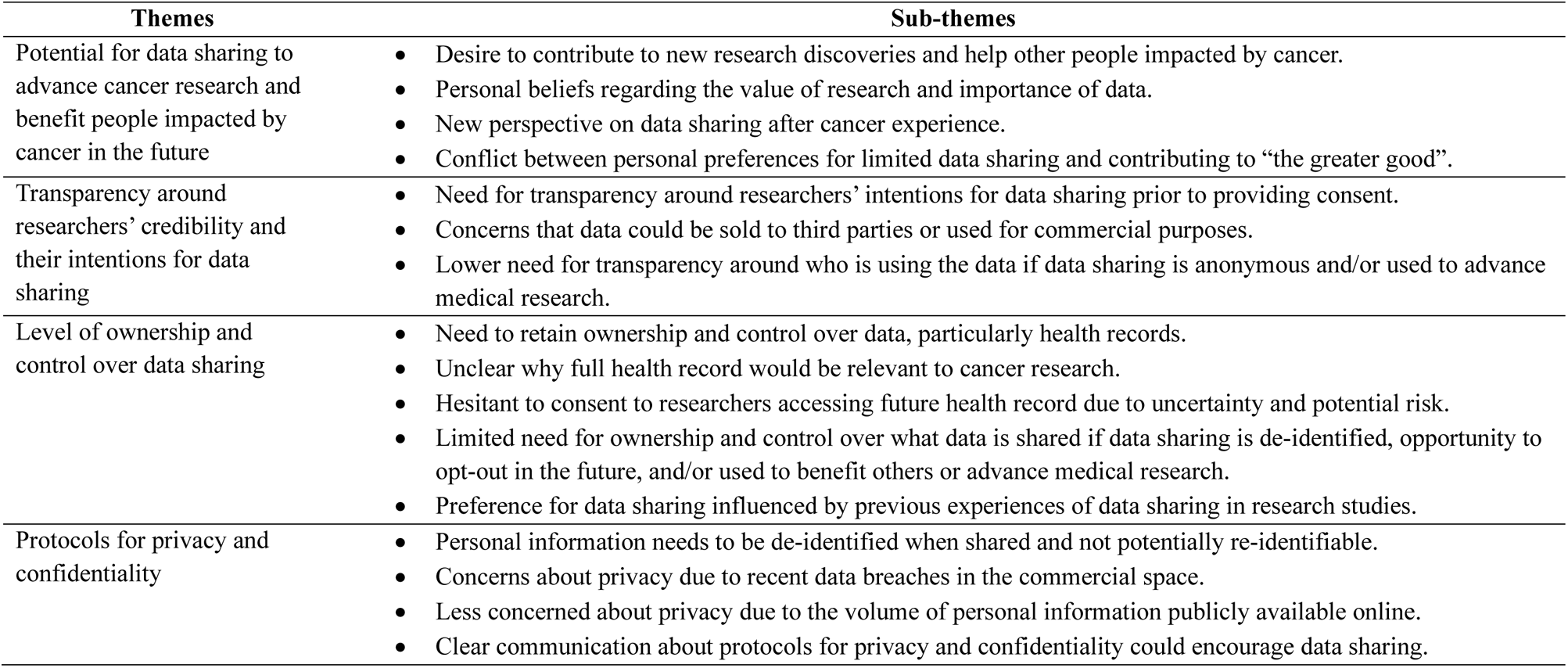
Themes and sub-themes identified in a thematic analysis of community members’ preferences for data access and sharing in cancer research and reasons underlying those preferences.

| Themes | Sub-themes |
| --- | --- |
| Potential for data sharing to advance cancer research and benefit people impacted by cancer in the future | • Desire to contribute to new research discoveries and help other people impacted by cancer.<br>• Personal beliefs regarding the value of research and importance of data.<br>• New perspective on data sharing after cancer experience.<br>• Conflict between personal preferences for limited data sharing and contributing to “the greater good”. |
| Transparency around researchers’ credibility and their intentions for data sharing | • Need for transparency around researchers’ intentions for data sharing prior to providing consent.<br>• Concerns that data could be sold to third parties or used for commercial purposes.<br>• Lower need for transparency around who is using the data if data sharing is anonymous and/or used to advance medical research. |
| Level of ownership and control over data sharing | • Need to retain ownership and control over data, particularly health records.<br>• Unclear why full health record would be relevant to cancer research.<br>• Hesitant to consent to researchers accessing future health record due to uncertainty and potential risk.<br>• Limited need for ownership and control over what data is shared if data sharing is de-identified, opportunity to opt-out in the future, and/or used to benefit others or advance medical research.<br>• Preference for data sharing influenced by previous experiences of data sharing in research studies. |
| Protocols for privacy and confidentiality | • Personal information needs to be de-identified when shared and not potentially re-identifiable.<br>• Concerns about privacy due to recent data breaches in the commercial space.<br>• Less concerned about privacy due to the volume of personal information publicly available online.<br>• Clear communication about protocols for privacy and confidentiality could encourage data sharing. |

### Theme 1: Potential for data sharing to advance cancer research and benefit people impacted by cancer in the future

This theme related to the purpose and outcomes of the proposed research and how that influenced participants’ willingness to share data. Participants expressed a willingness to share their self-report data and health records as part of a cancer research study if sharing this information would advance cancer research discoveries and help other people impacted by cancer. Participants emphasised the importance of research that “makes life better” and “makes a difference” for people affected by cancer in the future:

> “If my data and my experience and my records help someone research something that makes life better for people that are coming up behind me, I’m all for that.” *(P16, Workshop J, personal cancer diagnosis)*

> “I’ve been incredibly active in this space… I’m a consumer reviewer for [cancer research grants] … Would I give my data to big pharma to make an expensive drug that not many people can afford? No. Would I give my data to someone who is working towards the benefit of all? Yes. I love translational research. It makes a difference for the end users.” *(P42, Workshop A, carer)*

Personal beliefs regarding the value of research and importance of data encouraged data sharing:

> “[Data] is important, and it could help shape the way cancer is treated in the future if [researchers] have more data. The more data [researchers] have, the more knowledge they have about the situation.” *(P7, Workshop M, no personal experience of cancer)*

For some participants, their experience of cancer changed their perspective on data sharing. Having gained a better understanding of the importance of cancer research they indicated they would be more willing to share their personal information:

> “I probably used to be on the other end of the scale and didn’t really share much, particularly online. This cancer experience sort of flipped that for me. That’s my ‘why’… I would probably say yes to scenario 4 [future health records shared with other researchers for other projects]. I know based on my own experience with my son, the drug that saved him has never been used before (that we are aware of) in a child as young as him or his type of cancer.” *(P35, Workshop C, carer)*

Some participants expressed a conflict between their personal preference for limited data sharing and being more open to data sharing for “the greater good”:

> “I guess I have some uneasiness around [sharing] future health records… but when I heard [workshop participant] reiterate how important clinical trials are, I hesitated and I thought, maybe the greater good is more important.” (*P31, Workshop D, carer*)

> “As each [scenario] came up, my risk adverse nature kicked in. Thank you to the other participants – you’ve opened me up. I’m trying to let my walls down.” (*P32, Workshop D, carer*)

### Theme 2: Transparency around researchers’ credibility and their intentions for data sharing in cancer research

This theme addressed participants’ desire to know who is conducting the research and how their data would be used. Participants valued transparency around researchers’ intentions for data use prior to providing consent for data sharing:

> “I do like to know where the information is going. I think an important part of consent is being aware of [how the data will be used].” *(P35, Workshop C, carer)*

> “Every specific research [project that uses my data], I need to be aware of… What are the intentions of the project?” *(P21, Workshop H, no personal experience with cancer)*

Participants expressed concerns that their data could be sold to third parties or used for commercial purposes rather than “public good”. Participants indicated that the level of transparency around researchers’ credibility and intentions would influence their willingness to share data as part of cancer research:

> “I would need to be aware of who [the researchers] are. It could change my mind… Some people come to the table with terrific credibility. Others you may not know about. [You may be giving] the chance for someone to sell your information because we all know that happens. [I would] probably need to be aware of who they are, rather than just ‘researchers’, for me to be comfortable with that.” (*P4, Workshop O, carer*)

> “There are a number of researchers that are looking to privatize their ideas. They are looking for public money, but they are looking to commercialize their results. I’m a believer in public funds for public good. I wouldn’t give my data to any old researcher. My answer is quite nuanced. Is it public or private? What is the intent? Is the research going to be siloed? Is it going to see the light of day?” *(P42, Workshop A, carer)*

On the other hand, some participants indicated a lower need for transparency around data sharing, provided privacy and confidentiality were maintained or the data were used to advance medical research:

> “[I would be comfortable] as long as [my data] is still kind of contained, it’s just [accessible to] researchers, it is not out to the world. I don’t think I’d need to know the specific projects, but it would be nice to know when it is being used.” (*P34, Workshop C, carer*)

> “If I wasn’t made aware [of who was using the data and how], I’d be a little bit hesitant, but still be happy to give it if it’s medical research and it’s for a cure and I can help in any way.” (*P27, Workshop F, carer*)

### Theme 3: Level of ownership and control over data sharing in cancer research

This theme related to participants’ considerations about what data they would be willing to share as part of cancer research and their level of autonomy over this. Participants expressed a need to retain ownership and control over their data, particularly their health records. Participants also wanted the opportunity to choose their level of data sharing for each project:

> “I would look at everything on a case-by-case basis. I wouldn’t mind [researchers] using my health records but only if it’s for something that I considered fully before I opted in.” *(P15, Workshop J, personal cancer diagnosis and carer)*

> “I want to have ownership and control of my own data. I’m [name]. I’m not my body.” *(P28, Workshop F, no personal experience with cancer)*

> “I think you could [provide the option] ‘I’m willing to be contacted for future research projects’ [on the consent form]. You can give people the option to say, I do want to keep contributing, if there is something else in future, I just want to have a sphere of influence around that.” (*P32, Workshop D, carer*)

Some participants were unclear why all the information in their health record would be relevant to cancer research. This limited their willingness to share this data, specifically records not related to their cancer diagnosis:

> “I’ve opted out of my health record because I don’t feel it is anybody’s business knowing my vaccination status or any of my past history.” *(P25, Workshop G, personal cancer diagnosis*)

> “If it was going to cancer research, the only records that would be relevant in my mind is the cancer-related records. To throw in a lung condition … I think it would be more confusing. Not my full records. A lot of it wouldn’t be relevant at all.” (*P18, Workshop I, personal cancer diagnosis/ carer*)

Other participants were hesitant to grant researchers access to their future health information, mostly due to uncertainty around their future situation and the risk of personal and sensitive information being shared:

> “I guess I have some uneasiness around future health records because I don’t yet know what my future health issues will be. If it was something around fertility, that might be something quite private, something that is sensitive to me. Maybe I might not feel comfortable with that.” *(P31, Workshop D, carer)*

> “I don’t know what future health problems I’m going to have and how I’m going to react emotionally to them yet, so I don’t know if I want to put myself in a position to give out all my data for a future issue… [sharing] data I currently have now, very comfortable. But I don’t really know about the future if things will change.” (*P5, Workshop N, carer*)

Some participants expressed a limited need for ownership and control over their data if other conditions were met. These included de-identification, opportunity to opt-out, and the data being used “for good”:

> “If it is anonymous and being used to help or prevent something in the future, I personally don’t see anything wrong with it.” (*P41, Workshop A, carer*)

> “I’m pretty liberal with data sharing as long as there is a way to opt out should I need to.” (*P9, Workshop L, carer*)

> “If my data can help someone in the future not suffer as much or lead to a cure or better treatment, something that improves outlook for future patients, I’d be all for that.” *(P16, Workshop J, personal cancer diagnosis)*

Other participants indicated that their preference for data sharing was influenced by the degree of data sharing that they have previously consented to as part of a research study:

> “My health records have already been out in the world. I have been in clinical trials to help future research for cancer. So, whatever I can do to help.” (*P24, Workshop G, personal cancer diagnosis*)

> “I’ve participated in another study where I took medications and I consented to my medical records [being used] for that study only… that is okay, definitely not just for any project.” (*P19, Workshop I, carer*)

### Theme 4: Protocols for privacy and confidentiality of data sharing in cancer research

This theme addressed participants’ desire to know how their data would be shared with researchers and how it would be stored, protected and reported. Privacy and confidentiality were important to most participants, including their personal information being de-identified when shared and not potentially re-identifiable:

> “I wouldn’t want my name and date of birth and address and [phone] number being shared with everyone. If I’m just an entity in the system, a ‘male in their 40s that had this’ and ‘this was my treatment’, I’m totally fine.” *(P16, Workshop J, personal cancer diagnosis)*

> “As long as it [indicates] that my future health records will be shared in a non-identifiable manner… it has to be non-identifiable.” (*P33, Workshop D, carer*)

Some participants expressed concerns about privacy due to recent data breaches in the commercial space, emphasising the importance of ensuring secure data storage and sharing practices:

> “Is there any way the data can get out to the public? [This is] the one thing I need to ask in light of certain companies having problems with security or going down, like [company name]. How do we know the software protecting this information is safe enough?” *(P20, Workshop H, personal cancer diagnosis)*

> “The number one question is can it be hacked? Can it be scammed? If I’m giving you all my data, it’s lovely that you tell me it’s going to be fine, but I was with [company name] and [company name] and look what happened.” (*P39, Workshop B, personal cancer diagnosis*)

Others were less concerned about privacy due to the volume of personal information that is publicly available online:

> “If you don’t want to share your data for good, then the likelihood is that you will get your data hacked anyway, and your data may get lost. Data is just data.” *(P41, Workshop A, carer)*

> “As far as I’m concerned, people know everything about you anyway.” *(P14, Workshop K, no personal experience with cancer)*

> Participants agreed that clear communication with prospective research participants about protocols for privacy and confidentiality could encourage data sharing: “I think the important thing is to explain what checks and balances and protocols are in place around people’s private information, particularly in the health area. I think it would be important it was clearly pointed out in communication to the people involved in the survey. That would be the number one priority.” (*P8, Workshop M, personal cancer diagnosis*)

> “Unless you reassure them [prospective research participants], they are going to default to scenario one [no data sharing]. Not because they don’t believe in the usefulness of their information for research but because they don’t have the information about what’s in place [to protect their data]… You need to get your wording right up to do date about whatever the latest is for integrity in sharing data.” (*P39, Workshop B, personal cancer diagnosis*)

## Discussion

This study provides important information on the types of data that community members may be willing to share in cancer research, with whom, and for what purpose. In general, most community members are willing to share their self-report data and health records in a cancer research study, and many are willing to provide consent for this information to be shared with other researchers for other studies that they are made aware of. For example, community members were happy to consent to receiving information about future studies where their data could be used as they arose and to decide on data sharing on a case-by-case basis. In general, community members were less willing to share their future health records due to uncertainty around their future health and the potential for sensitive information to be disclosed. As this is a small qualitative study, we were unable to further explore preferences for data access and sharing in cancer research based on participant characteristics.

While we identified four themes underlying community members’ willingness to share their data in cancer research, the sub-themes presented within each theme reveal that these themes are intertwined and should be viewed together. For instance, if data sharing was anonymous and the research findings would benefit other people impacted by cancer in the future, then community members indicated a lower need for transparency around who was using their data and a lower need for ownership and control over the projects their data were used in. A reoccurring narrative throughout the themes was that clear communication around the potential benefits of the research, what data would be shared with whom and how this would be undertaken, and the protocols in place to ensure anonymity, could encourage community members to consent to data sharing in cancer research. This highlights the importance of designing study invitation materials with community members to optimise the readability, relevance, and acceptability of these materials (12).

The findings presented in this paper expand on previous studies investigating data sharing preferences among people diagnosed with cancer. Consistent with our findings, several studies from the United States have shown that cancer patients are generally willing to share their health data to improve care, both in general and for themselves (9–11). In those studies, patients have also reported a need for transparency and control regarding how, when, and why their data are used, and the importance of measures that protect their privacy and security (9–11). The current study demonstrates the applicability of these findings in the Australian setting, not only with cancer survivors, but also people caring for someone with cancer. A novel finding of this study is that the potential for research to benefit people affected by cancer in the future is a key motivation for consenting to data sharing. However, willingness to share data is often contingent on a combination of factors, such as the credibility of the research team and assurance of data anonymity, particularly in the context of increased cybersecurity concerns and breaches. By understanding these nuanced preferences, researchers can better design study protocols that align with the values and expectations of the community, thereby enhancing participation in cancer research.

Progressing data sharing in cancer research will require broad engagement with a range of stakeholders. In Australia, while the current data landscape includes many population-based datasets that are relevant to cancer research, there are several barriers to data sharing (18). These include siloed datasets with data custodians bound by restrictive legislation and approval processes for data access that can take years to negotiate and execute (18). These challenges are long-standing, with a systematic review published ten years ago reporting twenty potential barriers to data sharing in public health across technical, motivational, economic, political, legal, and ethical spheres (19). While addressing these barriers requires extensive consultation with, and commitment from, stakeholders and data custodians, findings from our study demonstrate community support for greater data sharing in cancer research and contribute to community engagement efforts to establish protocols for advancing data sharing in cancer research.

Based on our findings, there are several steps that researchers can take to support data sharing in cancer research, despite the system-level barriers. For example, researchers can emphasize on study recruitment materials or during debrief sessions how participants’ involvement in the research, including sharing data with other researchers for other projects, may benefit people impacted by cancer in the future and contribute to new medical discoveries, and how their anonymity will be maintained in data sharing and reporting. Researchers can also provide prospective participants with links to information about the research team and examples of how their previous work has been implemented in practice. If offering participants the option to consent to their data being used by other researchers for other projects, assure community members that they will be provided with information about each project and the researchers involved, and will have the to confirm consent or opt-out of sharing their data. Finally, if asking community members to consent to their full health record being used for a cancer research study, provide information on why non-cancer related records are relevant to the proposed research.

### Strengths and Limitations

This study included a diverse sample of community members, including people living in rural areas and those with English as a second language. A key limitation of this study is that participants were community members who had already consented to providing self-report data for a cancer research study. Therefore, findings from this study may not represent the views of community members in general, particularly those who prefer to not engage in research and those who experience geographical, technological, or language barriers to research participation; groups that are hard to reach but likely important to include in population-based cancer research studies. A large proportion of the workshop participants were female, however, there is limited evidence to suggest that gender influences data sharing preferences for health research in general (20,21). While the themes identified in this study were discussed by participants with and without a personal experience of cancer, the preferences for data access and sharing reported by the small number of people with no personal experience of cancer could change if they were to be diagnosed, or care for someone, with cancer (22).

Groupthink is a commonly cited limitation of group-based qualitative research, where members of a group seek cohesion and conformity in decision-making rather than diversity and different perspectives (23). While our workshops included group discussion around reasons underlying preferences for data access and sharing, the preferences reported by participants in nearly all the workshops were varied. Some participants did indicate that the group discussion challenged their own perspective on data sharing, however, few participants changed their preferred scenario after participating in the group discussion.

### Conclusions

Overall, these findings provide community support for improved data sharing in cancer research, a priority for advancing cancer control. Considerations underlying community members’ willingness to share data in cancer research studies included the potential for data sharing to advance medical discoveries and benefit others impacted by cancer in the future, transparency around researchers’ intentions for data sharing and their credibility, level of ownership and control over data sharing, and protocols for privacy and confidentiality. Findings provide several practical strategies for researchers to use when designing cancer research studies, including emphasizing on study invitation materials how participants’ involvement in the research will benefit people impacted by cancer in the future, explaining why access to participants’ full medical record is relevant for the proposed study, and providing opportunities for people to consent to being contacted about future research studies where their data could be used. Incorporating community preferences into the design and conduct of cancer research studies has the potential to optimise community participation and the applicability and translation of research findings into practice.

## Acknowledgments

The authors thank all individuals who participated in a workshop for this study.

## Data Availability Statement

The data that support the findings of this study are available on request from the corresponding author. The data are not publicly available due to privacy or ethical restrictions.

## Ethical Approval Statement

The study was approved by the Human Research Ethics Committee of the University of Southern Queensland (ETH2023-0140). All participants provided written informed consent prior to participation.

## Author contributions

**EAJ:** conceptualization, methodology, investigation, formal analysis, visualization, writing – original draft, writing – review and editing

**XEB:** project administration, data curation, investigation, formal analysis, writing – review and editing

**SKA:** conceptualization, methodology, investigation, formal analysis, writing – review and editing

**LZ:** project administration, writing – review and editing

**VLB:** supervision, writing – review and editing

**BCG:** conceptualization, funding acquisition, methodology, supervision, writing – review and editing

